# Anti-TRIM72 Autoantibodies in Idiopathic Inflammatory Myopathies

**DOI:** 10.1101/2025.07.23.25332079

**Authors:** Eugene Krustev, Tiara N. Safaei, Daniela Trejo-Zambrano, Lisa Christopher-Stine, Andrew Mammen, Julie J. Paik, Jemima Albayda, Christopher A. Mecoli, Brittany L Adler, Noah Weisleder, Wael Jarjour, Brendan Antiochos, Eleni Tiniakou

## Abstract

**Background and Purpose:** Tripartite motif-containing protein 72 (TRIM72) mediates tissue-repair following injury in several organs, including muscle and lung. Autoantibodies directed against TRIM72 (anti-TRIM72) have been identified in patients with idiopathic inflammatory myopathies (IIM) and disrupt TRIM72 function in vitro. We hypothesized that IIM patients positive for anti-TRIM72 antibodies would have a more severe clinical phenotype.

**Methods:** Sera from IIM patient (antisynthetase syndrome [ASyS], immune mediated necrotizing myopathy [IMNM], and dermatomyositis [DM]) and healthy controls (HC) were included. Anti-TRIM72 autoantibodies were tested using enzyme linked immunosorbent assay. Anti-TRIM72 testing was positive if value was >2 standard deviations above the mean for HC. Clinicodemographic features were identified through chart review and compared between anti-TRIM72 positive (anti-TRIM72[+]) and negative (anti-TRIM72[-]) groups.

**Results:** Anti-TRIM72 levels were significantly increased in patients with ASyS and IMNM when compared to patients with DM and healthy controls. Anti-TRIM72 levels were also increased in patients expressing anti-Jo-1, anti-PL7, anti-HMGCR, anti-SRP, and anti-MDA5. In ASyS, when anti-TRIM72(+) and anti-TRIM72(-) patients were compared, there were significantly more anti-TRIM72(+) ASyS patients with normal DLCO (>75%) when compared to anti-TRIM72(-); however, there were no differences in demographic features, CK levels or FVC. In anti-HMGCR(+) IMNM, anti-TRIM72(+) was associated with a lower proportion of females, as well as older age at time of diagnosis and at time of anti-TRIM72 testing; however, there was no significant difference in other clinicodemographic features in anti-HMGCR(+) IMNM patients when anti-TRIM72(+) and anti-TRIM72(-) groups were compared.

**Conclusions:** Anti-TRIM72 antibody titres are increased in patients with ASyS and IMNM. The presence of anti-TRIM72 antibodies was not associated with a more severe phenotype in ASyS or anti-HMGCR(+) IMNM, and there were more ASyS patients with normal DLCO in the anti-TRIM72(+) group.

## Introduction

The Idiopathic inflammatory myopathies (IIM) are a group of systemic autoimmune rheumatic diseases (SARD) that are characterized by autoimmune muscle involvement (Lundberg, 2021); however, extra-muscular manifestations are common in specific IIM subtypes. These extra-muscular manifestations include cutaneous, pulmonary, articular, vascular, oropharyngeal, gastrointestinal, and cardiac manifestations (Lundberg et al., 2021). The IIM subtypes are dermatomyositis (DM), antisynthetase syndrome (ASyS), immune-mediated necrotizing myopathy (IMNM), inclusion body myositis (IBM), overlap syndromes with myositis (OM), and polymyositis (PM) (Lundberg et al., 2021), with PM increasingly being recognized as a diagnosis of exclusion among experts in the field. Compared to the other SARD, the clinicoserologic phenotypes associated with specific IIM antibodies are relatively homogenous; therefore, antibody specificity is useful in predicting disease manifestations. The autoantibodies in IIM can be divided into two groups: myositis-specific antibodies (MSA), which are almost exclusively found in IIM and tend to correlate with a particular IIM subtype, and myositis-associated antibodies (MAA), which are found across the spectrum of SARD (Choi et al., 2023). Autoantibody positivity is common in IIM, with most studies showing that 60-70% of IIM patients are positive for either an MSA or MAA (Lundberg, 2021); however, many ‘seronegative’ patients likely have antibodies to currently unknown or not commonly tested autoantigens. As such, identifying novel autoantibodies in IIM can help identify unique phenotypes amongst subsets of IIM patients.

Tripartite motif-containing proteins (TRIM) are a family of E3 ubiquitin ligases that share a TRIM/RBCC (RING, B-box, coiled-coil) structure and have diverse biological functions in most cells, including the immune system, tumorigenesis, cell division, and plasma membrane repair (Vunjak & Versteeg, 2019). The TRIM family is of interest in IIM, as several TRIMs are target antigens for IIM-autoantibodies. Anti-Ro52/TRIM21 is frequently positive in IIM and other SARD, and anti-TIF1-γ (also known as p155/140, TRIM33) is an MSA. Furthermore, exploratory analyses have identified that several other autoantibodies targeting additional TRIMs are found in IIM patients (Megremis et al., 2021). These results suggest that the TRIMs are common antigen targets for SARD-associated autoantibodies, especially in IIM.

TRIM72 (also known as mitsugumin 53 or MG53) is expressed in striated muscle cells and is involved in sarcolemmal membrane repair following injury in skeletal and cardiac muscle (Cai, Masumiya, et al., 2009; Weisleder et al., 2012). Exogenous TRIM72 improves muscle pathology in pre-clinical mouse models of muscular dystrophy, suggesting a potential therapeutic benefit (Weisleder et al., 2012). TRIM72 also facilitates cellular and tissue repair in other organ systems, including the lung (Jia et al., 2014), brain (Y. Yao et al., 2016), liver (W. Yao et al., 2017) and eye (Chandler et al., 2019). In this role, MG53 integrates into the outer leaflet of the plasma membrane during repair patch formation, exposing it to the extracellular space (Cai, Weisleder, et al., 2009) and shedding of the protein into the circulation (Weisleder et al., 2012). In addition to its plasma membrane repair functions, TRIM72 promotes stem cell regeneration (Guan et al., 2019), decreases inflammation (Sermersheim et al., 2020), and has anti-viral effects in vivo (Kenney et al., 2021).

Given the role of TRIM72 in muscle physiology and the exposure of TRIM72 to the extracellular space following cell injury, autoantibodies directed against TRIM72 (anti-TRIM72) have been investigated in IIM by McElhanon and colleagues (McElhanon et al., 2020). In their study, 34.6% of DM and 21.6% of PM patients were positive for anti-TRIM72 antibodies, as were 57.9% of patients with elevated creatine kinase (CK). Anti-TRIM72 antibodies were also detectable in a mouse model of IIM. Furthermore, when myocytes were exposed to anti-TRIM72 antibodies in vitro, sarcolemmal membrane repair and resealing were impaired following experimental injury. There was a similar impairment in sarcolemmal membrane repair when myocytes were exposed to patient sera positive for anti-TRIM72 antibodies, and this effect was abrogated by depleting anti-TRIM72 from the sera. Based on these results, the authors concluded that anti-TRIM72 antibodies could directly hinder muscle repair in vivo.

Based on the role of TRIM72 in both muscle and lung repair mechanisms, the prominent occurrence of anti-TRIM72 antibodies in IIM patients, and the pathogenic effects of anti-TRIM72 antibodies on muscle repair in vitro, we hypothesized that we would identify anti-TRIM72 antibodies in our cohort of IIM patients and that these patients would have a more severe clinical phenotype, specifically higher muscle enzyme elevations and worse pulmonary function.

## Methods

### Patients and Healthy Controls

The study included patients evaluated at the Johns Hopkins Myositis Center (Baltimore, MD, USA) between 2002 and 2021. Patients with available serum samples and clinical data were randomly selected from the cohort with the goal of including 200 DM, 150 ASyS and 150 IMNM patients; however, some patients changed definitions after chart review or were excluded due to lack of follow up, uncertainty around diagnosis, or availability of serum samples. The study was approved by the Johns Hopkins Institutional Review Board (IRB00235256), and all participants provided informed consent. Demographic and clinical data (age at diagnosis, sex, ethnicity, mortality, CK levels, diffusing capacity of the lung for carbon monoxide [DLCO], forced vital capacity [FVC]) was collected through review of all available electronic medical records.

All IIM patients met the Bohan and Peter criteria for PM or DM (Bohan & Peter, 1975a, 1975b) and/or the 2017 European League Against Rheumatism/American College of Rheumatology classification criteria for adult and juvenile idiopathic inflammatory myopathies (Lundberg et al., 2017). Patients with immune-mediated necrotizing myopathy (IMNM) met the 2017 ENMC criteria for IMNM (Allenbach et al., 2018). Patients with ASyS met the 2024 ENMC criteria for ASyS (Stenzel et al., 2024). MSA (anti-Jo-1, anti-PL7, anti-PL12, anti-EJ, anti-OJ, anti-TIF1gamma, anti-Mi-2a, anti-NXP2, anti-Mi-2b, anti-MDA5, anti-SAE, anti-SRP) and MAA (anti-Ku, anti-PM/Scl-75, anti-PM/Scl-100) positivity was assessed by line blot (Euroline Autoimmune Inflammatory Myopathies 16Ag, EUROIMMUN, Lübeck, DE) using a positivity threshold of ≥15. Anti-HMGCR was tested by enzyme linked immunosorbent assay (ELISA; QUANTA Flash® HMGCR, Inova Diagnostics, San Diego, CA, USA) and a positivity threshold of ≥20 was used.

Additionally, where available, antibody testing that was done either for previous research or clinical purposes (immunoprecipitation or enzyme linked immunosorbent assay [ELISA]) was used to assign antibody status. Considering all available antibody tests, as well as clinical phenotype, patients were assigned an MSA or MAA.

Healthy control biospecimens were obtained as part of an ongoing IRB-approved study at the Johns Hopkins School of Medicine, where self-identified healthy volunteers aged 18 or older who were not pregnant and had no history of autoimmune disease, cancer, Lyme disease, or active infections such as HIV, tuberculosis, or hepatitis were recruited.

### Anti-TRIM72 ELISA

Anti-TRIM72 levels were tested on first available samples for each patient. A 96-well plate was coated with 200 ng/well (100 μL/well) of human TRIM72 protein (recombinant full length [477 amino acid] protein, expressed in E. coli; Origene, Rockville, MD, USA). The plate was incubated overnight at 4°C. Following incubation, the wells were washed twice with 300 µL PBS-T (1X PBS + 0.05% Tween-20). Blocking was performed by adding 100 µL of blocking agent (5% milk in PBS-T) per well, followed by incubation at room temperature (RT) for one hour while shaking. After blocking, the wells were washed, and 100 µL of IIM or HC sera diluted 1:500 in 1% milk-PBS-T was added per well, then incubated for two hours at RT while shaking. After incubation, the wells were washed three times with PBS-T. The secondary antibody (anti-human-HRP) was prepared at a 1:10,000 dilution in 1% milk-PBS-T and protected from light.

The wells were then incubated with 100 µL of the secondary antibody solution per well for one hour at RT with shaking. Following secondary antibody incubation, the wells were washed three times with PBS-T, followed by a final wash with PBS. SureBlue substrate (SeraCare, Gaithersburg, MD, USA) was brought to RT, and 100 µL was added per well. Color development was visually monitored for approximately 10 minutes by observing the standard curve. When the difference between the two lowest standard concentration became distinguishable, the reaction was stopped in 100 µL of 1N HCl per well. The absorbance was measured at 450 nm using a plate reader (Victor 3, Perkin Elmer, Waltham, MA, USA). Normalized values were calculated by dividing each value by the 1:400 dilution along the standard curve for each plate and are expressed as arbitrary units (AU). Anti-TRIM72 positivity (anti-TRIM72[+]) was defined as having anti-TRIM72 levels >2 standard deviations (SD) above the mean for healthy controls (normalized value > 0.9 AU), while patients who were below this cutoff were considered anti-TRIM72 negative (anti-TRIM72[-]).

## Statistical Analysis

All analyses and figures were generated using GraphPad Prism (GraphPad Software Inc., Boston, MA, USA). All continuous variables were compared using Mann-Whitney, Kruskal-Wallis, one-way analysis of variance (ANOVA) tests, and Pearson correlation, and are expressed as median with interquartile range (IQR). Categorial variables were compared using Fisher’s exact tests and are expressed as percentage positive (‘n’).

## Results

### Cohort Description

In total, there were 469 IIM patients included in this study (ASyS, n=182; IMNM, n=170; DM, n=117) and 67 HC (Table 1). Our IIM cohort had a median age of 53.6 at time of anti-TRIM72 testing, was 69.3% female, and 66.1% White race/ethnicity (Table 1). Median length of follow up was 5.9 years (IQR 1.9, 9.5) for the entire IIM cohort, 4.9 years (IQR 1.4, 9.6) for ASyS cohort, 5.5 years (IQR 0.7, 8.5) for IMNM cohort, and 7.6 years (IQR 5.4, 11.4) for DM cohort. A primary MSA or MAA was identified in 93.6% of our cohort, while 6.4% did not have an identifiable antibody (Table 1). Our HC cohort had a median age of 33.5 years, was 59.7% female, and 61.2% White race/ethnicity.

**Table 1:** Demographics and Antibody Positivity Across Whole Cohort.

|  | **Total IIM**<br>**(n=469)** | **ASyS**<br>**(n=182)** | **IMNM**<br>**(n=170)** | **DM**<br>**(n=117)** |
| --- | --- | --- | --- | --- |
| **Age (years)<sup>#</sup>,**<br>**median (IQR)** | 53.6 (43.1, 62.8) | 51.6 (41.5, 57.7) | 57.8 (45.1, 66.5) | 52.8 (44.0, 61.8) |
| **Female, n (%)** | 325 (69.3%) | 132 (72.5%) | 105 (61.8%) | 88 (75.2%) |
| **White, n (%)** | 310 (66.1%) | 103 (56.6%) | 112 (65.9%) | 95 (81.2%) |
| **Myositis**<br>**Antibodies, n (%)** |  |  |  |  |
| anti-Jo-1 | 125 (26.7%) | 125 (68.7%) | 0 (0.0%) | 0 (0.0%) |
| anti-PL-7 | 25 (5.3%) | 25 (13.7%) | 0 (0.0%) | 0 (0.0%) |
| anti-PL-12 | 21 (4.5%) | 21 (11.5%) | 0 (0.0%) | 0 (0.0%) |
| anti-OJ | 4 (0.9%) | 4 (2.2%) | 0 (0.0%) | 0 (0.0%) |
| anti-EJ | 5 (1.1%) | 5 (2.7%) | 0 (0.0%) | 0 (0.0%) |
| anti-Zo | 2 (0.4%) | 2 (1.1%) | 0 (0.0%) | 0 (0.0%) |
| anti-HMGCR | 100 (21.3%) | 0 (0.0%) | 100 (58.8%) | 0 (0.0%) |
| anti-SRP | 56 (11.9%) | 0 (0.0%) | 54 (32.9%) | 0 (0.0%) |
| anti-TIF1-γ | 35 (7.5%) | 0 (0.0%) | 0 (0.0%) | 35 (29.9%) |
| anti-NXP2 | 19 (4.1%) | 0 (0.0%) | 0 (0.0%) | 19 (16.2%) |
| anti-MDA5 | 13 (2.8%) | 0 (0.0%) | 0 (0.0%) | 13 (11.1%) |
| anti-Mi2 | 11 (2.3%) | 0 (0.0%) | 0 (0.0%) | 11 (9.4%) |
| anti-SAE | 6 (1.3%) | 0 (0.0%) | 0 (0.0%) | 6 (5.1%) |
| anti-PM/Scl-100/75 | 10 (2.1%) | 0 (0.0%) | 0 (0.0%) | 10 (8.5%) |
| anti-Ku | 2 (0.4%) | 0 (0.0%) | 1 (0.6%) | 1 (0.9%) |
| anti-RNP | 2 (0.4%) | 0 (0.0%) | 1 (0.6%) | 1 (0.9%) |
| AMA | 3 (0.6%) | 0 (0.0%) | 2 (1.2%) | 1 (0.9%) |
| Unknown | 30 (6.4%) | 0 (0.0%) | 10 (5.9%) | 20 (17.1%) |

<sup>#</sup>Age at first available blood draw used for anti-TRIM72 testing. **Abbreviations:** AMA, antimitochondrial antibodies; Anti-EJ, anti-glycyl tRNA synthetase; anti-HMGCR, anti-3-hydroxy-3-methylglutaryl-coenzyme A reductase; anti-Jo-1, anti-histidyl tRNA synthetase; anti-Ku, anti-Ku antigen; anti-MDA5, anti-melanoma differentiation-associated gene 5; anti-Mi2, anti-Mi2 antigen; anti-NXP2, anti-nuclear matrix protein 2; anti-OJ, anti-isoleucyl tRNA synthetase; anti-PL-12, anti-alanyl tRNA synthetase; anti-PL-7, anti-threonyl tRNA synthetase; anti-PM/Scl-100/75, anti-PM/Scl complex (100 and 75 kDa proteins); anti-RNP, anti-ribonucleoprotein; anti-SAE, anti-small ubiquitin-like modifier activating enzyme; anti-SRP, anti-signal recognition particle; anti-TIF1-γ, anti-transcription intermediary factor 1-gamma; anti-TRIM72, anti-
tripartite motif containing protein 72; anti-Zo, anti-phenylalanyl tRNA synthetase; ASyS, antisynthetase syndrome; DM, dermatomyositis; HC, healthy controls; IIM, idiopathic inflammatory myopathy; IMNM, immune mediated necrotizing myopathy; SD, standard deviation; unknown, no identifiable myositis-specific or myositis-associated antibodies.

### Anti-TRIM72 Positivity Across IIM Subtypes and Antibody Specificities

In total, 17.7% (n=83/469) of IIM patients in our cohort were positive for anti-TRIM72 antibodies (anti-TRIM72[+]), which was significantly greater than HC (n=0/67, 0.0%; p<0.0001; Table 2). When compared to HC, the proportion of anti-TRIM72(+) patients was significantly greater in ASyS (n=46/182, 25.3%; p<0.0001; Table 2) and IMNM (n=32/170, 18.8%; p<0.0001; Table 2). When compared to HC, there was no significant difference in the proportion of anti-TRIM72(+) DM patients (n=5/117, 4.3%; p=0.16; Table 2).

**Table 2:** Anti-TRIM72 Positivity Across IIM Subtypes and Antibody Specificities.

|  | Anti-TRIM72(+) <sup>#</sup> | Anti-TRIM72(-) | *p*<sup>##</sup> |
| --- | --- | --- | --- |
| HC | 0 (0.0%) | 67 (100.0%) | - |
| **Total IIM Cohort** | **83 (17.7%)** | **386 (82.3%)** | **<0.0001** |
| **ASyS** | **46 (25.3%)** | **136 (74.7%)** | **<0.0001** |
| **IMNM** | **32 (18.8%)** | **138 (81.2%)** | **<0001** |
| DM | 5 (4.3%) | 112 (95.7%) | 0.16 |
| Myositis Antibodies, n (%) |  |  |  |
| **anti-Jo-1** | **31 (24.8%)** | **94 (75.2%)** | **<0.0001** |
| **anti-PL-7** | **5 (20.0%)** | **20 (80.0%)** | **0.009** |
| **anti-PL-12** | **7 (33.3%)** | **14 (66.7%)** | **<0.0001** |
| anti-OJ | 1 (25.0%) | 3 (75.0%) | 0.056 |
| **anti-EJ** | **2 (40.0%)** | **3 (60.0%)** | **0.0039** |
| anti-Zo | 0 (0.0%) | 2 (100.0%) | >0.99 |
| **anti-HMGCR** | **21 (21.0%)** | **79 (79.0%)** | **<0.0001** |
| **anti-SRP** | **8 (14.3%)** | **48 (85.7%)** | **0.0014** |
| anti-TIF1-γ | 0 (0.00%) | 35 (100.0%) | >0.99 |
| anti-NXP2 | 0 (0.00%) | 19 (100.0%) | >0.99 |
| **anti-MDA5** | **3 (23.1%)** | **10 (76.9%)** | **0.018** |
| anti-Mi2 | 0 (0.0%) | 11 (100.0%) | >0.99 |
| anti-SAE | 0 (0.0%) | 6 (100.0%) | >0.99 |
| anti-PM/Scl-100/75 | 0 (0.0%) | 10 (100.0%) | >0.99 |
| **anti-Ku** | **1 (50.0%)** | **1 (50.0%)** | **0.029** |
| **anti-RNP** | **1 (50.0%)** | **1 (50.0%)** | **0.029** |
| AMA | 0 (0.0%) | 3 (100.0%) | >0.99 |
| **Unknown** | **3 (10.0%)** | **27 (90.0%)** | **0.028** |

<sup>#</sup>Anti-TRIM72(+) defined as > 2 SD above the mean for HC (0.9 AU). <sup>##</sup>p value when compared to HC. Significant comparisons noted in **bold**. Abbreviations: AMA, antimitochondrial antibodies; Anti-EJ, anti-glycyl tRNA synthetase; anti-HMGCR, anti-3-hydroxy-3-methylglutaryl-coenzyme A reductase; anti-Jo-1, anti-histidyl tRNA synthetase; anti-Ku, anti-Ku antigen; anti-MDA5, anti-melanoma differentiation-associated gene 5; anti-Mi2, anti-Mi2 antigen; anti-NXP2, anti-nuclear matrix protein 2; anti-OJ, anti-isoleucyl tRNA synthetase; anti-PL-12, anti-alanyl tRNA synthetase; anti-PL-7, anti-threonyl tRNA synthetase; anti-PM/Scl-100/75, anti-PM/Scl complex (100 and 75 kDa proteins); anti-RNP, anti-ribonucleoprotein; anti-SAE, anti-small ubiquitin-like modifier activating enzyme; anti-SRP, anti-signal recognition particle; anti-TIF1-γ, anti-transcription intermediary factor 1-gamma; anti-TRIM72, anti-tripartite motif containing protein 72; anti-Zo, anti-phenylalanyl tRNA synthetase; ASyS, antisynthetase syndrome; DM, dermatomyositis; HC, healthy controls; IIM, idiopathic inflammatory myopathy; IMNM, immune
mediated necrotizing myopathy; SD, standard deviation; unknown, no identifiable myositis-specific or myositis-associated antibodies.

When compared to HC, the proportion of anti-TRIM72(+) was significantly greater in patients positive for the ASyS-associated autoantibodies anti-Jo-1 (n=31/125, 24.8%; p<0.0001; Table 2), anti-PL7 (n=5/25, 20.0%; p=0.009; Table 2), anti-PL12 (n=7/21, 33.3%; p<0.0001; Table 2) and anti-EJ (n=2/5, 40.0%; p=0.0039; Table 2). The IMNM-associated autoantibodies anti-HMGCR (n=21/100, 21.0%; p<0.0001; Table 2) and anti-SRP (n=8/56, 14.3%; p=0.0012; Table 2) also had greater co-occurrence with anti-TRIM72 when compared to HC. Compared to HC, the proportion of anti-TRIM72(+) was also significantly increased in patients with the DM-associated antibody anti-MDA5 (n=3/13, 23.1%; p=0.018; Table 2). The MAA autoantibodies, anti-Ku (n=1/2, 50.0%; p=0.029; Table 2) and anti-RNP (n=1/2, 50.0%; p=0.029; Table 2), which are frequently associated with overlap syndromes, also had higher rates of anti-TRIM72(+) when compared to HC. Our seronegative group, who did not express a known MSA or MAA had three patients who were anti-TRIM72(+) (3/30; 10.0%; p=0.028; Table 2), which was greater than HC. Anti-TRIM72(+) did not differ from HC in the other MSA and MAA tested (p’s>0.05; Table 2).

### Anti-TRIM72 Titres Across IIM Subtypes and Autoantibody Specificities

When anti-TRIM72 levels were compared to HC, anti-TRIM72 levels were significantly greater in ASyS (0.44 vs 0.17; p<0.0001; Figure 1A) and IMNM (0.41 vs 0.17; p<0.0001; Figure 1A). Compared to HC, there was no significant difference in anti-TRIM72 levels in DM (0.23 vs 0.17; p=0.86; Figure 1A).

**Figure 1:**
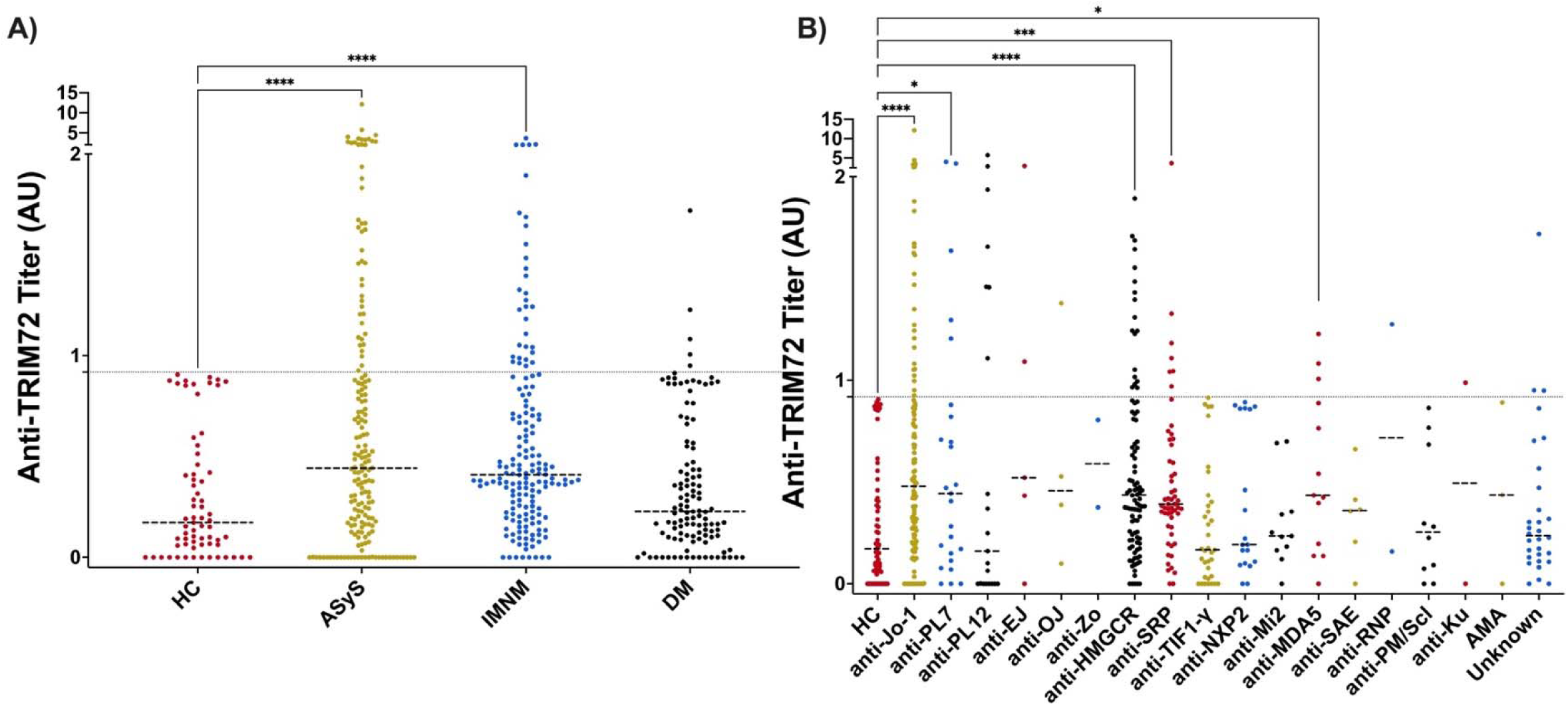
Anti Anti-TRIM72 Antibody Levels Across IIM Subtypes and Antibody Specificities. A) Anti-TRIM72 levels (AU) in HC, ASyS , IMNM and DM. B) Anti-TRIM72 levels in patients positive for MSA or MAA. Thick dotted line denotes median for each group. Thin dotted line denotes >2 SD cutoff (0.9 AU) above HC. Abbreviations: AMA, antimitochondrial antibodies; Anti-EJ, anti-glycyl tRNA synthetase; anti-HMGCR, anti-3-hydroxy-3-methylglutaryl-coenzyme A reductase; anti-Jo-1, anti-histidyl tRNA synthetase; anti-Ku, anti-Ku antigen; anti-MDA5, anti-melanoma differentiation-associated gene 5; anti-Mi2, anti-Mi2 antigen; anti-NXP2, anti-nuclear matrix protein 2; anti-OJ, anti-isoleucyl tRNA synthetase; anti-PL-12, anti-alanyl tRNA synthetase; anti-PL-7, anti-threonyl tRNA synthetase; anti-PM/Scl-100/75, anti-PM/Scl complex (100 and 75 kDa proteins); anti-RNP, anti-ribonucleoprotein; anti-SAE, anti-small ubiquitin-like modifier activating enzyme; anti-SRP, anti-signal recognition particle; anti-TIF1-γ, anti-transcription intermediary factor 1-gamma; anti-TRIM72, anti-tripartite motif containing protein 72; anti-Zo, anti-phenylalanyl tRNA synthetase; AU, arbitrary units; ASyS, antisynthetase syndrome; DM, dermatomyositis; HC, healthy controls; IIM, idiopathic inflammatory myopathy; IMNM, immune mediated necrotizing myopathy; unknown, no identifiable myositis-specific or myositis-associated antibodies.*p<0.05, ***p<0.001, ****p<0.0001.

When compared to HC, anti-TRIM72 titres were significantly increased in patients positive for anti-Jo-1 (0.48 vs 0.17 ; p<0.0001; Figure 1B), anti-HMGCR (0.44 vs 0.17; p<0.0001; Figure 1B), anti-SRP (0.39 vs 0.17; p=0.0005; Figure 1B), anti-MDA5 (0.43 vs 0.17; p=0.018; Figure 1B) and anti-PL7 (0.44 vs 0.17; p=0.017; Figure 1B). Compared to HC, there was no significant difference in anti-TRIM72 levels in patients positive for any other MSA or MAA tested (p’s>0.05; Figure 1B).

### Anti-TRIM72 clinicodemographic associations in ASyS

Given the association of anti-TRIM72(+) in ASyS and increased co-occurrence with multiple ASyS-associated autoantibodies, we compared demographic and clinical features in ASyS patients with and without anti-TRIM72 antibodies.

Anti-TRM72(+) ASyS patients were 71.7% female, which was not significantly different from those who were anti-TRIM72(-) (72.8%; p=0.89; Table 3). There was no significant difference in the proportion of patients who were of White race/ethnicity when compared between anti-TRIM72(+) and anti-TRIM72(-) ASyS patients (54.4% vs 57.4%; p=0.73; Table 3). There was no significant difference in age at diagnosis in anti-TRIM72(+) and anti-TRIM72(-) ASyS patients (50.7 years vs 48.1 years; p=0.22; Table 3), nor was there a difference in age at anti-TRIM72 testing (53.6 vs 51.1; p=0.26; Table 3).

**Table 3:** Clinicodemographic Features Associated with Anti-TRIM72 in ASyS Patients.

|  | Total Cohort<br>n=182 | Anti-TRIM72(+)<br>n=46 | Anti-TRIM72(-)<br>n=136 | p-value<sup>#</sup> |
| --- | --- | --- | --- | --- |
| Female, n (%) | 132 (72.5%) | 33 (71.7%) | 99 (72.8%) | 0.89 |
| White, n (%) | 103 (56.6%) | 25 (54.4%) | 78 (57.4%) | 0.73 |
| Age at diagnosis, median (IQR) | 48.4 (38.5, 55.1) | 50.7 (44.0, 57.9) | 48.1 (38.1, 54.7) | 0.22 |
| Age at anti-TRIM72 testing, median (IQR) | 51.6 (41.5, 57.7) | 53.6 (46.5, 60.3) | 51.1 (40.6, 57.4) | 0.26 |
| Highest recorded CK (U/L), median (IQR) | 1969.0 (600.0, 6236.0) | 1401.0 (261.5, 6618.0) | 2005.0 (682.8, 6195.5) | 0.71 |
| Normal CK (<200U/L), n (%) | 23 (12.6%) | 8 (17.4%) | 15 (11.0%) | 0.26 |
| CK at time of anti-TRIM72 test (U/L), median (IQR) | 388.5 (150.8, 1395.0) | 412.0 (161.0, 1826.0) | 386.0 (144.0, 1352.0) | 0.71 |
| Lowest recorded DLCO (ml/min/mmHg), median (IQR) | 11.3 (8.3, 15.6) | 9.9 (6.3, 15.0) | 12.0 (8.6, 15.7) | 0.15 |
| Lowest recorded DLCO % predicted, median (IQR) | 51.0 (36.0, 67.0) | 42.0 (29.0, 75.5) | 54.0 (37.9, 66.8) | 0.24 |
| **Normal DLCO (>75%), n (%)** | **33/177 (18.6%)** | **14/43 (32.6%)** | **19/134 (14.2%)** | **0.0071\*\*** |
| Lowest recorded FVC (L), median (IQR) | 2.2 (1.7, 2.9) | 2.1 (1.6, 3.0) | 2.3 (1.7, 2.8) | 0.82 |
| Lowest recorded FVC % predicted, median (IQR) | 62.3 (48.9, 76.8) | 64.5 (44.9, 79.8) | 62.0 (49.8, 75.7) | >0.99 |
| Normal FVC (>80% predicted), n (%) | 64/181 (35.4%) | 18/46 (39.1%) | 46/135 (34.1%) | 0.54 |
| Deceased, n (%) | 37 (20.3%) | 10 (21.7%) | 27 (19.9%) | 0.83 |

<sup>#</sup>p value calculated for anti-TRIM72(+) vs anti-TRIM72(-) groups. Significant comparisons noted in **bold**. Abbreviations: ASyS, anti-synthetase syndrome; CK, creatine kinase; DLCO, diffusing capacity of the lung for carbon monoxide; FVC, forced vital capacity; TRIM72, tripartite motif containing 72.

When highest recorded CK levels were compared between anti-TRIM72(+) and anti-TRIM72(-) ASyS patients, there was no significant difference between groups (1401.0 U/L vs 2005.0 U/L; p=0.71; Table 2). We also compared ASyS patients with persistently normal CK (<200.0 U/L) versus those who had CK elevations; 17.4% anti-TRIM72(+) ASyS patients had persistently normal CK while 11.0% anti-TRIM72(-) ASyS patients failed to mount an elevated CK, which was not significantly different (p=0.24; Table 3). There was no significant correlation between anti-TRIM72 antibody levels and highest recorded CK concentrations (Pearson correlation, r = 0.011; 95% CI: –0.136 to 0.157; p = 0.887; Supplemental Figure 1). Additional analyses of highest recorded CK (log_10_CK and grouping based on the severity of CK elevation) did not result in any significant differences between anti-TRIM72(+) and anti-TRIM72(-) ASyS patients (data not shown).

Since both CK and antibody levels can fluctuate over time, we also compared differences in CK near time of anti-TRIM72 testing. There were 110 ASyS patients (anti-TRIM72[+], n=89; anti-TRIM72[-], n=21) who had a CK tested within six weeks of the date that their anti-TRIM72 sample was drawn. CK levels were not significantly different between anti-TRIM72(+) and anti-TRIM72(-) groups (412.0 U/L vs 386.0 U/L; p=0.71; Table 3) and anti-TRIM72 antibody levels were not significantly correlated with serum CK levels measured within six weeks of testing (Pearson correlation, r = 0.083; 95% CI: -0.11 to 0.27; p = 0.39; Supplemental Figure 1).

When lowest recorded DLCO was assessed in ASyS patients, there was no significant difference between anti-TRIM72(+) and anti-TRIM72(-) patients in absolute DLCO (9.9 mL/min/mmHg vs 12.0 mL/min/mmHg; p = 0.15; Table 3), nor DLCO % predicted (42.0 vs 54.0; p=0.24; Table 3). When ASyS patients were grouped based on those with a persistently normal DLCO (>=75% predicted) and those who had a reduced DLCO (<75% predicted), there were significantly more patients in the anti-TRIM72(+) group with persistently normal DLCO when compared to anti-TRIM72(-) (32.6% vs 14.2%; p = 0.007; Table 3). Anti-TRIM72 antibody levels were not significantly correlated with DLCO % predicted (Pearson correlation, r = 0.036; 95% CI: -0.11 to 0.18; p = 0.63; Supplemental Figure 1).

When lowest recorded FVC was assessed is ASyS patients, there was no significant difference between anti-TRIM72(+) and anti-TRIM72(-) patients in absolute FVC (2.1 L vs 2.3 L; p=0.82; Table 3), nor FVC % predicted (64.5 vs 62.0; p>0.99; Table 3). When patients were grouped based on normal FVC (>=80% predicted) and abnormal FVC (<80% predicted), there was no significant difference in the proportion of patients with normal FVC between anti-TRIM72(+) and anti-TRIM72(-) ASyS patients (39.1% vs 34.1%; p=0.54; Table 3). No significant correlation was observed between anti-TRIM72 antibody levels and lowest recorded FVC % predicted (Pearson correlation, r = 0.052; 95% CI: -0.096 to 0.20; p = 0.49; Supplemental Figure 1). Additional analyses where patients were grouped based on the severity of the worst FVC % predicted were also non-significant (data not shown).

There was no significant difference in the proportion of ASyS patients who died during the follow up period between anti-TRIM72(+) and anti-TRIM72(-) patients (21.7% vs 19.9%; p=0.83; Table 3).

We also assessed ASyS patients who were anti-Jo-1 positive as this was the largest antibody specific group amongst the ASyS patients and was significantly associated with anti-TRIM72(+). When comparing those who were anti-TRIM72(+) (n=31) to those who were anti-TRIM72(-) (n=94) we found no significant differences in age at diagnosis (50.7 vs 48.2; p=0.17), proportion of female patients (71.0% vs 76.6%; p=0.63), highest recorded CK (2424.0 U/L vs 2396.0 U/L; p=0.89), lowest recorded DLCO % predicted (46.0 vs 55.0; p = 0.25), or lowest recorded FVC % predicted (65.0 vs 63.0; p=0.96).

### Anti-TRIM72 clinicodemographic associations in anti-HMGCR IMNM

Given the prevalence of anti-TRIM72 antibodies in patients with IMNM and anti-HMGCR antibodies, we compared anti-HMGCR IMNM patients who were anti-TRIM72(+) with those who were anti-TRIM72(-).

There were significantly fewer female anti-HMGCR IMNM patients who were anti-TRIM72(+) when compared to the anti-TRIM72(-) group (33.3% vs 60.8%; p=0.029; Table 4). There was no significant difference in the proportion of anti-HMGCR IMNM patients who were of White race/ethnicity when compared between anti-TRIM72(+) and anti-TRIM72(-) groups (71.4% vs 74.7%; p=0.78; Table 4). When age at diagnosis was compared, anti-HMGCR IMNM anti-TRIM72(+) patients were older at the time of diagnosis when compared to their anti-TRIM72(-) counterparts (66.1 vs 58.3; p=0.041; Table 4). Similarly, when age at anti-TRIM72 testing was compared between groups, anti-TRIM72(+) patients were significantly older when compared to anti-TRIM72(-) in anti-HMGCR IMNM (69.1 vs 61.1; p=0.0043; Table 4).

**Table 4:** Clinicodemographic Features Associated with Anti-TRIM72 in Anti-HMGCR(+) IMNM.

|  | **Total Cohort**<br>**n=100** | **Anti-TRIM72(+)**<br>**n=21** | **Anti-TRIM72(-)**<br>**n=79** | **p-value<sup>#</sup>** |
| --- | --- | --- | --- | --- |
| **Female, n (%)** | **55 (55.0%)** | **7 (33.3%)** | **48 (60.8%)** | **0.029** |
| White, n (%) | 74 (74.0%) | 15 (71.4%) | 59 (74.7%) | 0.78 |
| **Age at diagnosis,**<br>**median (IQR)** | **59.3 (52.0, 67.1)** | **66.1 (50.3, 72.6)** | **58.3 (51.8, 65.6)** | **0.041** |
| **Age at anti-TRIM72**<br>**testing, median (IQR)** | **62.3 (54.9, 68.9)** | **69.1 (62.2, 73.3)** | **61.1 (54.3, 66.0)** | **0.0043** |
| Statin Exposed, n (%) | 82/99<sup>##</sup> (83.17%) | 18/20<sup>##</sup> (90.00%) | 64 (81.0%) | 0.51 |
| Worst CK, median<br>(IQR) | 5385.0 (2523.0,<br>9132.0) | 5168.0 (3652.5,<br>6928.0) | 5540.0 (2323.0,<br>9341.0) | 0.94 |
| Worst CK >5000, n (%) | 54 (54.0%) | 12 (57.1%) | 42 (53.2%) | 0.81 |
| CK at time of anti-TRIM72 test (U/L),<br>median (IQR) | 1890.0 (508.0,<br>5017.0) | 1899.0 (1062.0,<br>5454.0) | 1843.0 (359.3,<br>4815.0) | 0.73 |
| Deceased, n (%) | 13 (13.0%) | 1 (4.8%) | 12 (15.2%) | 0.29 |

<sup>#</sup>p value calculated for anti-TRIM72(+) vs anti-TRIM72(-) groups. <sup>##</sup>One patient excluded due to missing data. Abbreviations: CK, creatine kinase; IMNM, immune mediated necrotizing myopathy; TRIM72, tripartite motif containing 72.

Given that increased age and male sex are associated with statin-exposed anti-HMGCR IMNM when compared to statin-naïve disease, we also compared statin exposure history between anti-TRIM72(+) patients (n=20, one patient excluded due to missing data) and anti-TRIM72 (-) patients, and there was no significant difference in the proportion of statin-exposed patients between groups (90.0% vs 81.0%; p=0.51; Table 4).

When highest recorded CK levels were assessed in anti-HMGCR IMNM patients, there was no significant difference between anti-TRIM72(+) and anti-TRIM72(-) (5168.0 U/L vs 5540.0 U/L; p=0.94; Table 4). When patients were grouped based on CK elevation >5000 U/L, there was no significant difference in the proportion of patients in the anti-TRIM72(+) group when compared to anti-TRIM72(-) patients (57.1% vs 53.2%; p=0.81; Table 4). When we compared CK levels that were tested within six weeks of anti-TRIM72 testing, there was no significant difference between anti-TRIM72(+) and anti-TRIM72(-) patients (1899.0 vs 1843.0; p=0.73; Table 4). There was no significant correlation between anti-TRIM72 antibody levels and the highest recorded serum CK values (Pearson correlation, r = 0.099; 95% CI: -0.010 to 0.29; p = 0.33; Supplemental Figure 1), nor with CK recorded within six weeks of anti-TRIM72 testing (Pearson correlation, r = -0.046; 95% CI: -0.29 to 0.21; p = 0.72; Supplemental Figure 1).

There was no significant difference in the proportion of anti-HMGCR IMNM patients who died during the follow up period between anti-TRIM72(+) and anti-TRIM72(-) patients (4.8% vs 15.2%; p=0.29; Table 4).

## Discussion

The results of this study show that anti-TRIM72 antibodies are frequently positive in IIM patients with ASyS and IMNM. Previous work has shown that anti-TRIM72 was increased in DM patients (McElhanon et al., 2020); however, we did not see an increased signal in this IIM subtype in our cohort, and DM anti-TRIM72 levels were comparable to HC. One likely reason for this difference is that we subtyped patients into ASyS, IMNM, and DM, while McElhanon and colleagues grouped patients into DM and PM. Based on these groupings, it is possible that some of these DM patients would have been in our ASyS group. McElhanon and colleagues also saw anti-TRIM72(+) in their PM group (McElhanon et al., 2020). While we did not classify patients as PM in our cohort, many of the PM patients from their study would have likely been split between our IMNM and ASyS groups. Together, our study and theirs show that anti-TRIM72 antibodies are frequently found in multiple subtypes of IIM patients.

Interestingly, anti-TRIM72 antibodies were more frequently observed in ASyS and IMNM when compared to DM in our study. Muscle histology from ASyS and IMNM tends to have more necrosis when compared to DM (Selva-O’Callaghan et al., 2018), and necrotic muscle cells may result in greater release of cellular contents (Kamiya et al., 2023). Moreover, the membrane repair process itself can result in exposure of TRIM72 to the extracellular space (Demonbreun et al., 2016) or its release into the extracellular space (Weisleder et al., 2012; Williams et al., 2024). The release of cellular contents in the setting of myocyte membrane repair and necrosis may allow more TRIM72 exposure to immune cells and autoantibody formation. Another possibility is that anti-TRIM72 antibodies may contribute to myocyte necrosis by impairing membrane repair in ASyS and IMNM. Although we did not specifically compare differences in muscle histology, future studies should assess differences in histopathology between anti-TRIM72 (+) and anti-TRIM72 (-) patients as this may help us better understand how these autoantibodies develop and potentially contribute to disease pathogenesis.

Anti-TRIM72 antibodies also co-occurred with several MSA and MAA, specifically with ASyS-associated and IMNM-associated antibodies. Our results show that anti-TRIM72 antibodies appear in IIM patients with similar frequency as several common MSA and MAA. Anti-TRIM72(+) was also increased in patients positive for the DM-specific antibody anti-MDA5 and the overlap-associated antibodies anti-Ku and anti-RNP; however, the number of patients in each of these groups were low and should be interpreted with caution. One research priority in IIM is the identification of novel antibodies to fill the ‘seronegative gap’ in patients with a clinical diagnosis of IIM but who are not positive for a known or tested antibody (Choi et al., 2023). In our study, only three patients in our seronegative group were anti-TRIM72(+), and anti-TRIM72 occurred more frequently with other known MSA and MAA; therefore, based on our results, anti-TRIM72 does not appear to be a strong candidate for identifying ‘seronegative’ patients.

TRIM72 is involved in sarcolemmal repair and interacts with dysferlin and caveolin-3 to aid in vesicular trafficking to the site of injury during patch formation (Cai, Weisleder, et al., 2009). In vitro, purified anti-TRIM72 and patient serum containing anti-TRIM72 antibodies have been shown to impair sarcolemmal membrane repair in muscle injury models (McElhanon et al., 2020). As such, we hypothesized that anti-TRIM72(+) IIM patients would have more severe muscle damage and subsequently higher CK levels; however, anti-TRIM72 was not associated with higher CK levels in ASyS or anti-HMGCR(+) IMNM in our study. Based on these results, we cannot conclude that anti-TRIM72 is associated with clinically worse muscle damage in IIM. This could be in part due to the elevated levels of TRIM72 expression in skeletal muscle from IIM patients (McElhanon et al., 2020) that could increase sarcolemmal membrane repair, compensate for the effects of anti-TRIM72 antibodies and minimize the leak of CK from skeletal muscle.

TRIM72 also facilitates tissue repair in extramuscular tissues, specifically the lungs, which is particularly interesting in IIM, as pulmonary involvement in the form of ILD is quite common – especially in ASyS (Ghanbar & Danoff, 2024). When lung function in anti-TRIM72(+) and anti-TRIM72(-) ASyS patients was assessed by comparing impairments in both lung volume (FVC) and gas transfer (DLCO), there were no significant differences between groups in quantitative measures of lung function; however, there were significantly more anti-TRIM72(+) ASyS patients with persistently normal DLCO when compared to the anti-TRIM72(-) group. To our knowledge, this is the first study to look at anti-TRIM72 antibodies in ASyS-associated pulmonary disease. Based on these results, we cannot conclude that anti-TRIM72 is associated with clinically worse lung disease in IIM.

Although it is difficult to make any firm conclusions if anti-TRIM72 antibodies play a role in disease pathogenesis based on these results, the fact that anti-TRIM72(+) patients were more likely to have normal DLCO, may suggest a less severe clinical phenotype. Although most pathogenic autoantibodies impair antigen function, there are examples of autoantibodies that activate their target antigens in human disease, such as thyroid-stimulating immunoglobulins in Grave’s disease (Smith & Hall, 1974); therefore, it is possible that anti-TRIM72 binding promotes TRIM72 function in vivo, which may facilitate tissue repair; however, this would contradict previous in vitro studies showing an impairment in sarcolemmal resealing in the presence of anti-TRIM72 antibodies (McElhanon et al., 2020). Another possibility is that patients with more effective tissue repair mechanisms may have increased TRIM72 expression at the site of injury, which could then result in increased autoantibody formation. It is interesting to note that TRIM72 protein expression is greater in skeletal muscle when compared to lungs (Kim et al., 2014), which might explain why we saw a difference in pulmonary function but not in CK levels. Future studies are needed to determine how anti-TRIM72 affects tissue repair in vivo and the mechanisms that result in the development of these autoantibodies.

Strengths of our study include a large cohort size encompassing the major IIM subtypes that we suspected would be anti-TRIM72(+), as well as a long follow up period. A limitation of our study is that all patients were seen at a single center; however, given the nature of our myositis clinic, patients come from many parts of the United States and globally, resulting in a diverse patient population. Given the retrospective nature of our study, we were limited to selecting outcome measures available for all of our patients to ensure consistency. Although CK is the most frequently measured biomarker for muscle involvement in IIM, there can be significant discordance across patients, with some patients who are very weak having relatively mild elevations in CK and vice versa. We also could not assess differences in skeletal muscle function testing or magnetic resonance imaging (MRI) scores of muscle structure across all patients. Similarly, we included the worst FVC and DLCO as these were available for all patients; however, other measures of pulmonary disease, such as specific radiographic patterns of ILD, may provide further clues into the differences in lung disease between anti-TRIM72(+) and anti-TRIM72(-) patients. Future studies should aim to incorporate these measures of muscle and lung disease, as well as treatment responses, to help better understand the potential pathogenic effects of anti-TRIM72 antibodies and how they might contribute to disease severity. Moreover, the examination of anti-TRIM72 antibodies longitudinally should be informative, especially if there is a compensatory increase in the expression of TRIM72 in a particular organ.

In summary, this study confirms previous findings that anti-TRIM72 is frequently positive in IIM; however, based on our results above, anti-TRIM72 antibodies did not result in a more severe clinical phenotype, which was our original hypothesis. Conversely, anti-TRIM72 positivity may confer a milder phenotype with a greater likelihood of having preserved alveolar gas transfer. Future studies are needed to characterize how anti-TRIM72 antibodies develop and affect tissue repair in vivo.

## Data Availability

All data produced in the present work are contained in the manuscript

---

**Supplemental Figure 1.**
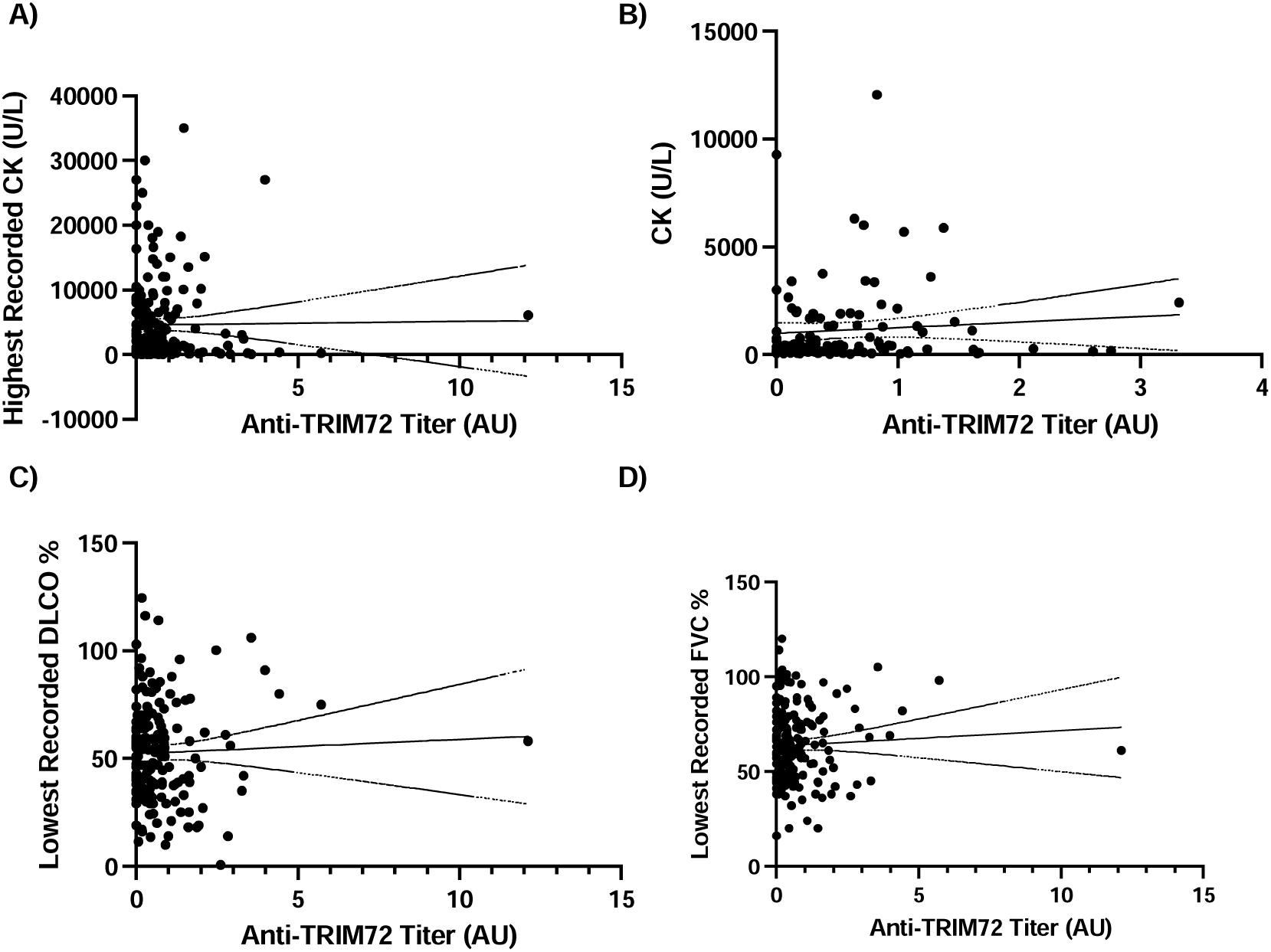
: Simple Linear Regression Comparing Anti-Trim72 Titers with Clinical Features in ASyS. A) Anti-TRIM72 titers are not correlated with highest recorded CK (U/L). B) Anti-TRIM72 titers are not correlated with CK (U/L) drawn within six weeks of anti-TRIM72 testing. C) Anti-TRIM72 titers are not correlated with lowest recorded DLCO % predicted. D) Anti-TRIM72 titers are not correlated with lowest recorded FVC % predicted. Abbreviations: ASyS, anti-synthetase syndrome; CK, creatine kinase; DLCO, diffusing capacity of the lung for carbon monoxide; FVC, forced vital capacity; TRIM72, tripartite motif containing 72.

**Supplemental Figure 2.**
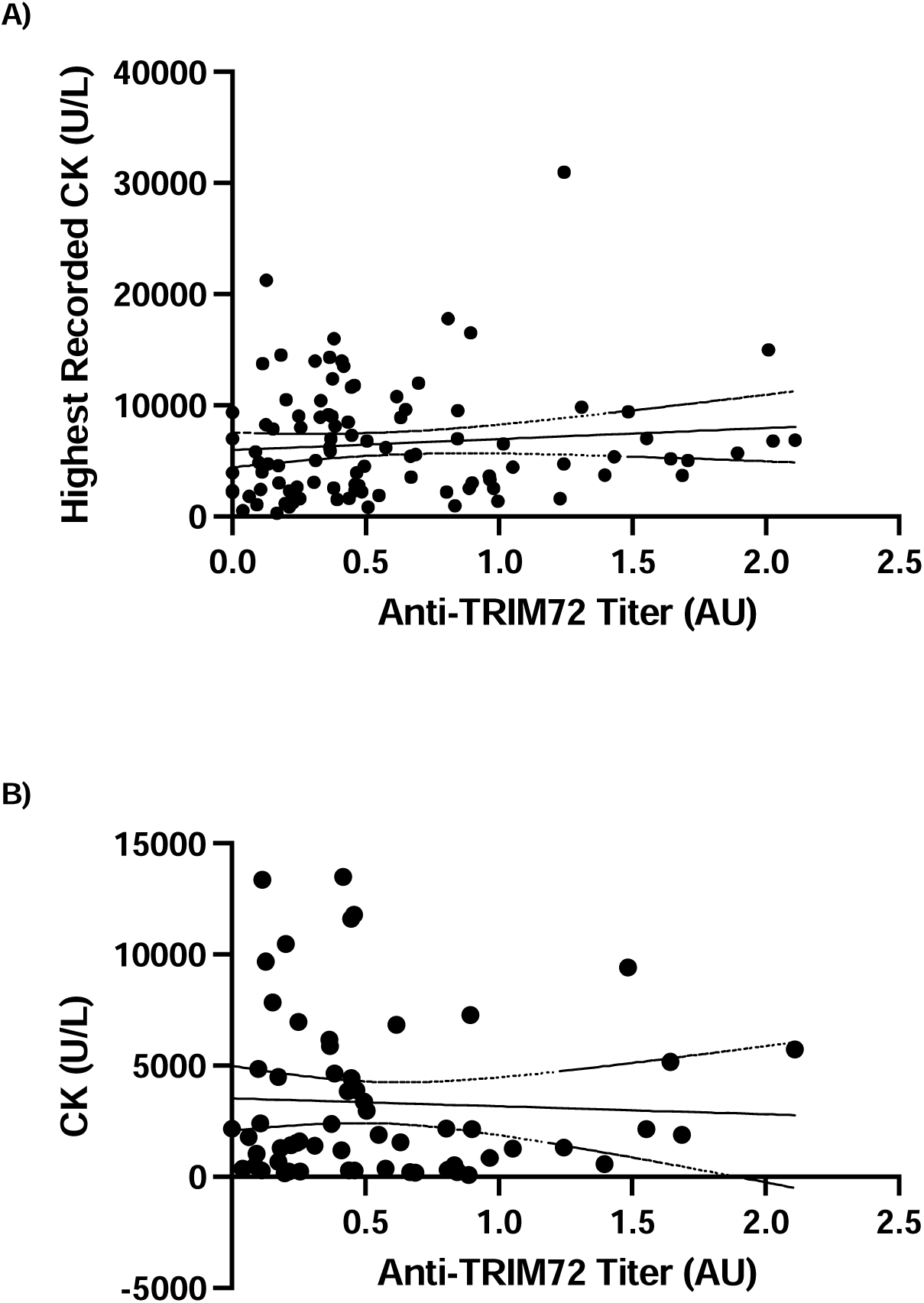
: Simple Linear Regression Comparing Anti-Trim72 Titers with CK Levels in Anti-HMGCR IMNM. A) Anti-TRIM72 are not correlated with highest recorded CK (U/L). B) Anti-TRIM72 titers are not correlated with CK (U/L) drawn within six weeks of anti-TRIM72 testing. Abbreviations: CK, creatine kinase; IMNM, immune mediated necrotizing myopathy; TRIM72, tripartite motif containing 72.

